# Negative SARS-CoV-2 PCR or rapid antigen test result and the subsequent risk of being infectious: a mathematical simulation study

**DOI:** 10.1101/2021.03.12.21253440

**Authors:** Ralf Krumkamp, Benno Kreuels, Veronika K. Jaeger, Jürgen May, Rafael Mikolajczyk, André Karch

## Abstract

**Background:** A considerable proportion of SARS-CoV-2 transmission occurs from asymptomatic and pre-symptomatic cases. Therefore, different polymerase chain reaction (PCR)- or rapid antigen test (RAT)-based approaches are being discussed and applied to identify infectious cases that would have gone undetected (e.g., in nursing homes). In this article, we provide a framework to estimate the time-dependent risk of being infectious after a negative SARS-CoV-2 test and we simulate the number of expected cases over time in populations of individuals who initially tested negative.

**Methods:** A Monte Carlo approach is used to simulate infections that occurred over a one-week period in populations with 1,000 individuals following a negative SARS-Cov-2 test. Parameters representing the application of PCR tests or RATs are utilized, and SARS-CoV-2 7-day incidences between 25 and 200 per 100,000 people are considered. Simulation results are compared to case numbers predicted via a mathematical equation.

**Results:** The simulations showed a linear increase in cases over time in populations of individuals who initially tested SARS-CoV-2 negative. The different false negative rates of PCR tests and RATs have a strong impact on the number of simulated cases. The simulated and the mathematically predicted case numbers were comparable. However, Monte Carlo simulations highlight that, due to random effects, infectious cases can exceed predicted case numbers even shortly after a test was conducted.

**Conclusions:** The analysis demonstrates that the number of infectious cases in a population can be effectively reduced by the screening of asymptomatic individuals. However, the time since the negative test and the underlying SARS-CoV-2 incidence are critical parameters in determining the observed subsequent number of cases in tested populations.

## Background

A considerable proportion of individuals with SARS-CoV-2 infection are free of symptoms or only show very mild symptoms. However, transmission can occur from both asymptomatic and pre-symptomatic cases [1,2]. Different PCR (polymerase chain reaction) or rapid antigen test (RAT)-based approaches are currently being considered or have already been implemented to identify cases that would have otherwise gone undetected (for example, to protect clinically vulnerable individuals in high-infection risk settings like nursing homes, to reduce unnecessary quarantine of non-infectious people, or to lift social contacts restrictions e.g. to permit care home visiting [3]).

For such measures to be effective, laboratory tests would ideally be done in real-time, as the test result reflects the current state of infectiousness of an individual. Since this is not always possible, especially for PCR analyses, tests done within a certain time frame are accepted. In travel restrictions, this time frame is usually 48h before travel [4]. In contrast, RAT results are available within 20–30 min. However, these tests have a lower sensitivity than PCR tests [5]. The time between sample taking and critical event (e.g., air travel) is a crucial parameter in determining the current risk of being infectious. An individual who has tested negative may be in the latent period of infection at sampling and could progress to an infectious state immediately thereafter. As time since testing increases, people with a previous negative test results will have the same risk of being infectious as the underlying population.

In this article, we provide a framework to estimate the time-dependent risk of being infectious after a negative PCR test or RAT, and we simulate the number of expected cases over time in populations of individuals who initially tested negative.

## Methods

We simulate infections that occur over a one-week period in a group of people who are SARS-CoV-2 negative at *t*_*0*_. For the simulations we assume that the population is entirely susceptible with no immunity, individuals are not suspected of being SARS-CoV-2 positive, and new cases are expected to occur homogeneously over time (i.e., unclustered). The basic simulation models are set up as follows: (i) a SARS-CoV-2 negative population is established; (ii) within this group, infectious cases are determined according to the predefined 7-day incidence following a binomial distribution; (iii) the time when infectiousness starts is allocated for each case; and (iv) the duration of infectiousness is allocated for each case, so that (v) the number of infectious cases within the population over time can be summarised.

PCR-based and RAT-based testing strategies are simulated. Parameters are taken from the literature as summarised in Table 1. Most SARS-CoV-2 cases transmit the infection within the first three days of their infectious period, and the majority of cases are non-infectious after 5 to 9 days [1,6]. For the simulations, an average infectious period of 96 hours (Gaussian distribution; standard deviation [SD]: 10) is assumed. PCR tests and RATs differ in their false negative rate (FNR), which defines the proportion of positive cases receiving a negative test result. FNRs depend on the specificity of a test and the accuracy of the test implementation. Reported FNRs for SARS-CoV-2 PCR tests vary greatly [7] and 3% was used as base case value. For RATs, the World Health Organization recommends minimum performance requirements of 80% sensitivity [8], which was assumed for calculations of the FNR in the RAT scenarios. Viral loads rise quickly at the beginning of the infectious period. However, in the first 12 hours after a PCR test is able to detect an infection, high circle threshold (ct) values are observed (i.e., low viral loads) and cases are assumed to be non-infectious during this time interval [9,10]. Hence, we determined the first 12 hours (Gaussian distribution; mean = 12, SD: 1) after a case would be PCR positive to be non-infectious. For both tests, simulations with reported 7-day incidences of 25, 50, 100 and 200 per 100,000 people were calculated. The 7-day incidences, as reported by surveillance systems, primarily capture symptomatic SARS-CoV-2 cases. However, only 35% of all SARS-CoV-2 cases are expected to be symptomatic and the remaining 65% show very mild or no symptoms [11]. To estimate the actual number of infectious cases within the simulated populations, the assumed incidences (as reported by surveillance systems) are divided by the proportion of symptomatic cases. Since absolute case numbers in the model population will be small, the Monte Carlo method was applied to show stochastic effects on the occurrence of cases. Simulations are repeated 2,000 times using parameter distributions as outlined above. To summarise the Monte Carlo results the number of infectious cases over time in the simulation runs were tabulated.

**Table 1:** Parameters applied in the Monte Carlo simulations.

| Parameter | Value | Reference |
| --- | --- | --- |
| Reported SARS-CoV-2 incidences | 25, 50, 100 & 200 /100,000 per 7 days |  |
| Proportion symptomatic infections | 0.35 | [11] |
| FNR for PCR test | 3% | [7] |
| FNR for RAT | 20% | [8] |
| Infectious period | Mean = 96 hours (SD: 10) | [1,6] |
| Time between PCR positivity and infectiousness | Mean = 12 hours (SD: 1) | [9,10] |
| Group size | 1,000 |  |
| Model runs | 2,000 |  |

Abbreviations: FNR, False negative rate; PCR, polymerase chain reaction; RAT, rapid antigen test; SD, standard deviation;

The expected number of infectious cases over time (*C*_*t*_) within a population which had a negative test result for SARS-CoV-2 can be described mathematically. In each scenario, *C*_*t*_ is based on the actual daily incidence of infectious cases per 100,000 people (*I*, representing the true number of infectious cases in a population), and the rate of prevalent cases corresponds to *I* times the infectious period (*ip*, measured in days). In a population of size *N*, where all individuals are SARS-CoV-2 test negative at *t*_*0*_, new cases start to emerge successively. The number of cases increases linearly over time, until first cases become non-infectious. Thus, as soon as first cases complete their infectious periods, the number of newly emerging cases and the number of cases becoming non-infectious is balanced. Thereafter, the previously negative population shows case frequencies as expected by the underlying incidence of infection. Assuming homogeneous case occurrence over time and taking the FNR of the test into consideration, the number of cases in a previously negative population at a particular day (defined in *t*) after negative testing can be estimated by

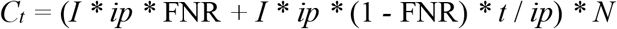

if *t* ≤ *ip. I * ip \** FNR describes the number of cases not detected by the test, and *I * ip \** (1 *-* FNR) *\* t* / *ip* represents the linear case-increase over time. Since the calculation is based on the incidence rate, it has to be multiplied by *N* to estimate the total number of cases.

The case numbers from the Monte Carlo simulations are compared to the mathematically predicted case numbers calculated by the equation derived above. To do this, the mean numbers of cases over time for the respective Monte Carlo scenarios is calculated. Furthermore, the equation is applied to calculate the expected case numbers over time using simulation parameters and both results are displayed using line graphs. All calculations were done in R version 4.0.3 [12].

## Results

To visualise the occurrence of infectious cases within a previously SARS-CoV-2 negative population, a simple baseline simulation was established (Figure 1). The simulation represents a population of 1,000 individuals, assuming a reported 7-day incidence of 200 cases per 100,000 people and a mean infectious period of 96 hours (SD = 10). The horizontal black lines in Figure 1 show infectious periods of cases that emerged over one week. In total, 7 infectious cases appeared during the simulation. The first case occurred 3 hours after the simulation start and the highest number of cases was observed at the end of day 7, where 5 individuals were infectious simultaneously.

**Figure 1:**
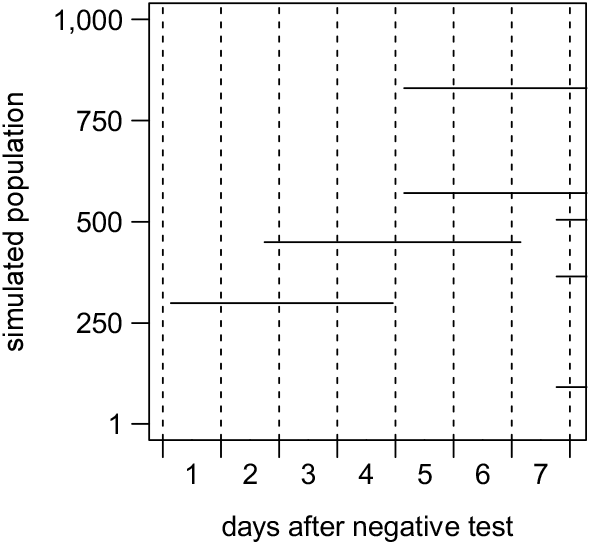
Example of a single baseline model, showing simulated infectious periods in a population of 1,000 individuals over one week.

To capture stochastic effects, the Monte Carlo method was employed and simulations were repeated 2,000 times based on the scenarios outlined above. Figure 2 summarises the results of the Monte Carlo simulations using area graphs. Graphs in the first row show simulation results of the PCR test and those in the second row of the RAT strategy. The graphs summarise simulation results based on different reported 7-day incidences over time. The proportion of simulations and the respective number of cases over time is indicated by colours.

**Figure 2:**
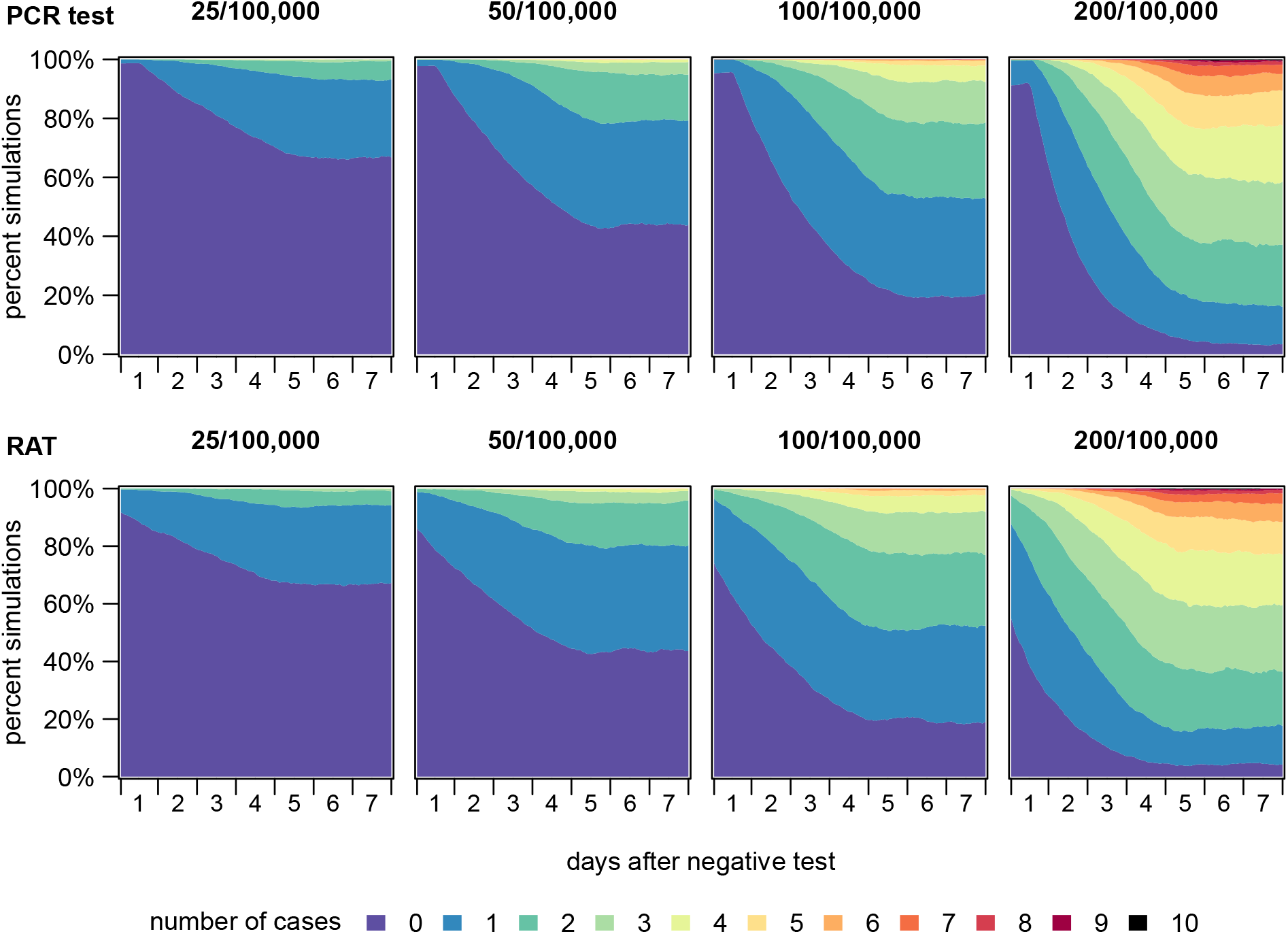
Percentages of simulations with different case numbers calculated using Monte Carlo method, considering different scenarios in a population of 1,000 individuals. Abbreviations: PCR, polymerase chain reaction; RAT, rapid antigen test.

Due to the assumed FNR of 3% for PCR tests, infectious cases occurred as early as at the start of some simulations. No infectious cases were observed at the start in 99% of simulations when the reported 7-day incidence was 25/100,000, in 98% of simulations when incidence was 50/100,000, in 95% when incidence was 100/100,000, and in 91% when incidence was 200/100,000. After one day (24 hours), no cases occurred at the different incidence levels in 94%, 88%, 80% and 64% of simulations, and after two days (48 hours) in 85%, 71%, 54% and 28% of the simulations. After 4.5 days (108 hours), the simulated case numbers started to stabilise and no further case increase was observed. Eventually, by the end of the simulation period (7 days) no infectious cases occurred in 67%, 43%, 20% and 4% of the simulations, which represents distributions in non-selected populations. Throughout time, multiple infections were likely to occur in scenarios with higher incidences. At a reported 7-day incidence of 25/100,000, a prevalence of 4 simultaneous cases was never exceeded. However, after 80 hours, 51 hours and 14 hours, more than 4 cases were observed in simulations based on 7-day incidences of 50, 100 and 200 cases per 100,000, respectively.

The FNR of the RAT was set to 20%, resulting in a lower proportion of simulations without any infectious cases at the simulation start. No infectious cases were observed at the start of the simulations in 92%, 86%, 74% and 55% of the simulations at incidence levels of 25, 50, 100 and 200 per 100,000, respectively. After one day (24 hours), no cases occurred in 85%, 72%, 53% and 28% of simulations, and after two days (48 hours) in 79%, 61%, 38% and 15% of the simulations, respectively. After 4 days (96 hours), the estimated number of cases started to stabilise and no infectious cases had occurred by the end of the simulation period in 67%, 44%, 19% and 4% of the simulations, which is comparable to the numbers calculated by the PCR scenarios. Similar to the PCR test, in simulations based on a reported 7-day incidence of 25/100,000, no simulation showed more than 4 cases; however, at 7-day incidences of 50,100 and 200/100,000, after 48 hours, 16 hours, 1 hour, respectively more than 4 cases were observed.

To compare case numbers simulated by Monte Carlo method and estimated by the mathematical equation derived above, the mean number of cases over time per scenario was calculated. The equation was applied to calculate the expected case numbers over time using the respective simulation parameters. Figure 3 shows calculated case numbers from the PCR test (first plot) and the RAT scenarios (second plot). The mean case numbers from the simulations are shown by the black lines, and the estimated case numbers calculated with the equation by the red lines. Case numbers calculated by both methods overlap. In both graphs, a linear case increase over a period of 4 days (the mean infectious period) is observed, after which the number of cases remain stable. These plateaus correspond to the expected number of infectious cases in a population of 1,000 people considering the respective incidences (dashed lines). Usually, PCR tests turn positive as soon as 12 hours before onset of infectiousness, which is why case numbers remain constant at the beginning of the simulation periods in the first graph.

**Figure 3:**
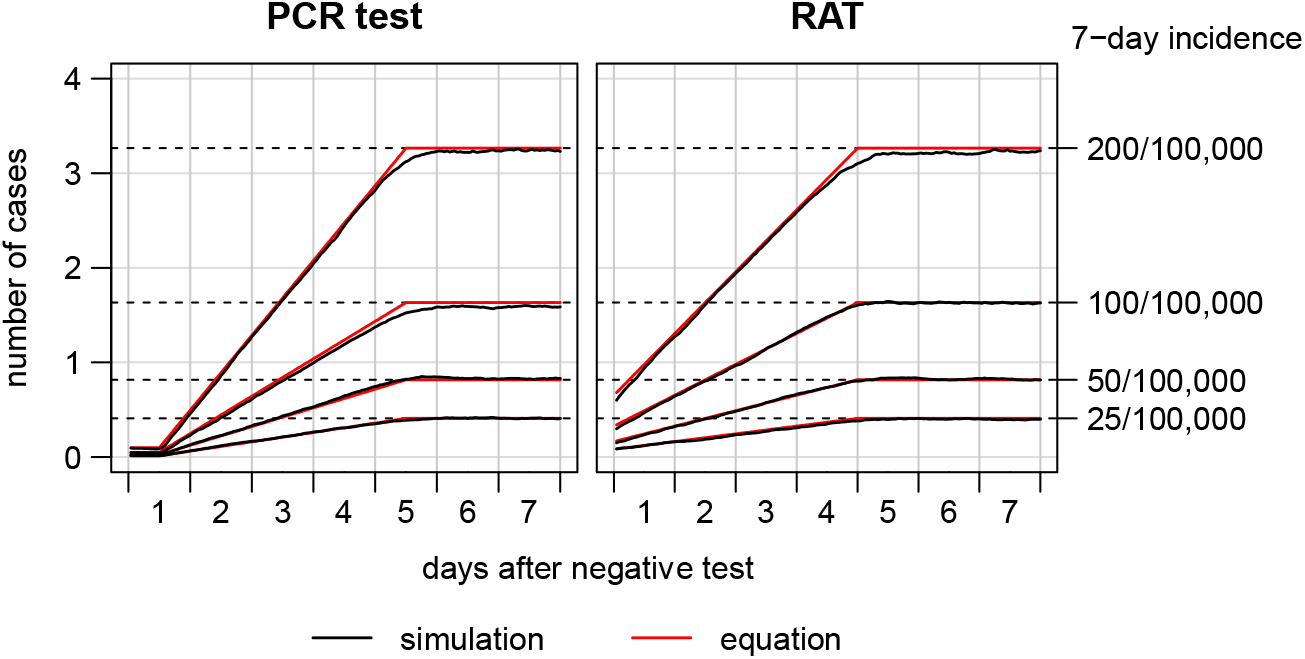
Number of cases over time in a group of 1,000 people averaged over the Monte Carlo simulation scenarios (red lines) and estimated by the equation as derived in the method section (black lines). Abbreviations: PCR, polymerase chain reaction; RAT, rapid antigen test.

## Discussion

SARS-CoV-2 can be transmitted from cases before they develop symptoms and some infectious cases do not develop any symptoms at all [1,2]. These infection characteristics require strategies beyond symptom-based screening in order to reduce pre- and asymptomatic transmission, responsible for a substantial number of SARS-CoV-2 cases.

Our analysis shows that testing asymptomatic individuals with PCR tests or RATs can reduce the number of infectious cases within populations effectively. We provide an easily applicable mathematical formula to estimate the expected case numbers over time using the disease incidence, the average infectious period and the time that has passed since a test was administered. However, the Monte Carlo simulations performed in our analysis highlight how, due to random effects, infectious cases can exceed these expected numbers even shortly after all individuals in the population had tested negative. In a fraction of simulated scenarios, single infectious cases occurred right after the simulation start; in high-incidence simulations, even multiple cases occurred. These results are important for infection control because they demonstrate that, while testing can be used to effectively select populations with a low number of infectious cases, especially at high incidences, it is still likely that infectious cases will start to emerge immediately after the population had tested negative.

PCR tests are superior to RATs in terms of their FNR [5] and their ability to identify cases even before they are infectious [9,10]. In contrast, RATs can be applied in non-laboratory settings and the results are available within 30 minutes. For PCR tests, times between testing and reporting of results of 24 to 48 hours are reported. Hence, the higher risk of falsely diagnosing a case as negative through the use of RATs is balanced by the immediate availability of the test result. If the high FNR of RATs is to be compensated for, it is important that the test is carried out immediately before a critical event takes place. The higher FNR still bears the risk of missing infectious cases, but the immediate application reduces the number of cases which may emerge before the critical event takes place.

Simplifying assumptions that were made to illustrate principles of SARS-CoV-2 testing should be considered when interpreting the results. Random case occurrence was assumed; however, SARS-CoV-2 is reported to spread in clusters and via super-spreading events [13]. Thus, in the case that testing is performed on a group of people where a superspreading event occurred (e.g., residents and personnel of a nursing facility with an ongoing SARS-CoV-2 outbreak), the prevalence of infectious cases would be considerably higher compared to the numbers reported here. The proportion of infectious cases without or with only mild symptoms was set to 65%. However, a correct estimate of this proportion is subject to several methodological limitations that make interpretation of the reported frequencies of symptoms among SARS-CoV-2 cases difficult [14]. Additionally, we assume that the remaining 35% of cases are recognized by a health care system and they contribute to the observed incidence at population level. These figures are very context-dependent and subject to current testing strategies. However, for many health care systems, lower reporting rates should be assumed. In the literature, there are conflicting reports about infectious periods in asymptomatic, mild, moderate or severe symptomatic cases as well as among different age-groups [15]. However, the infectious period is a parameter central in determining the number of cases and the temporal dependence in the occurrence of new cases.

SARS-CoV-2 control strategies based on diagnostics for asymptomatic individuals are suggested and designed for different purposes. A model of SARS-CoV-2 outbreaks in long-term care facilities evaluated the ability of different testing strategies to identify ongoing transmission early. The authors highlight that expanding surveillance beyond symptom-based screening could allow for earlier outbreak detection; however, testing strategies must consider available testing capacities [16]. Another study modelled the effect of surveillance testing to control SARS-CoV-2 transmission, concluding that asymptomatic individuals should be considered in testing strategies. Effective surveillance depends largely on the frequency of testing and the speed of reporting, and is only marginally improved by high test sensitivity [11]. These studies highlight the effect of testing asymptomatic individuals to control the spread of SARS-CoV-2, which is also supported by our study. With the current analysis we provide the rationale for estimating the risk of being infectious after a negative diagnostic test. Our analysis highlights the temporal dynamics of SARS-CoV-2 infections after a negative test within a theoretical population. We show that PCR tests or RATs can be used to select populations with a reduced number of SARS-CoV-2 cases. However, the parameters representing time since a negative test was conducted and the underlying SARS-CoV-2 incidence in a population are critical in determining the expected number of cases in test negative groups of people. Thus, especially in high-incidence scenarios, additional infection control measures are still needed to reduce transmission risk from undetected infectious cases.

## Data Availability

The R-code of this simulation study is available from the corresponding author, RaK, upon request.

## Acknowledgements

We thank Lydia Rautman for textual revision of the manuscript.

## Ethical approvals

Ethics approval was not required for this mathematical simulation study.

## Data availability

The R-code of this simulation study is available from the corresponding author, RaK, upon request.

